# A prospective, single-center, randomized phase 2 trial of etoposide in severe COVID-19

**DOI:** 10.1101/2023.06.05.23290969

**Authors:** Meredith Halpin, Adam Lerner, Manish Sagar, Praveen Govender, Bhavesh Shah, Janice Weinberg, Shayna Sarosiek, J. Mark Sloan

**Affiliations:** Boston University Chobanian & Avedisian School of Medicine Department of Medicine Section of Hematology and Oncology; Boston University Chobanian & Avedisian School of Medicine Department of Medicine Section of Infectious Disease; Boston University Chobanian & Avedisian School of Medicine Department of Medicine Section of Pulmonology; Boston University School of Public Health; Dana Farber Cancer Institute

## Abstract

The systemic inflammatory response seen in patients with severe COVID-19 shares many similarities with the changes observed in hemophagocytic lymphohistiocytosis (HLH); a disease characterized by excessive immune activation. Many patients with severe COVID qualify for a diagnosis of HLH. Etoposide, an inhibitor of topoisomerase II is used to control inflammation in HLH. This randomized, open-label, single center phase II trial attempted to determine whether etoposide can be used to blunt the inflammatory response in severe COVID. This trial was closed early after eight patients were randomized. This underpowered trial did not meet its primary endpoint of improvement in pulmonary status by two categories on an 8 point ordinal scale of respiratory function. There were not significant differences in secondary outcomes including overall survival at 30 days, cumulative incidence of grade 2 through 4 adverse events during hospitalization, duration of hospitalization, duration of ventilation and improvement in oxygenation or paO2/FIO2 ratio or improvement in inflammatory markers associated with cytokine storm. A high rate of grade 3 myelosuppression was noted in this critically ill population despite dose reduction, a toxicity which will limit future attempts to explore the utility of etoposide for virally-driven cytokine storm or HLH.

## Background

The initial wave of COVID-19 swept through the Northeast United States in the spring of 2020. At that time Boston Medical Center (BMC), a safety net hospital, had the largest proportional load of patients with COVID-19 infection in the city of Boston. In April 2020 seven out of 10 patients admitted to BMC patients had COVID-19 (Staff et al., n.d.). An alarming report from New York in April 2020 described the mortality for mechanically ventilated COVID-19 patients to be between 25 and 97%(Richardson et al., 2020). Reliable information on the risks and benefits of dexamethasone, remdesivir, therapeutic anticoagulation, tocilizumab and other interventions were not available during the first year of the pandemic as multi-center clinical trials were initiated, accrued and analyzed (Gottlieb et al., 2022; RECOVERY Collaborative Group et al., 2021; REMAP-CAP Investigators et al., 2021; Rosas et al., 2021). In the interim, these medications and others were used outside the context of clinical trials either as hospital policy or on a case-by-case basis.

It was noted early in the course of the pandemic that patients with severe disease often had evidence of a profound systemic inflammatory response (Ruan et al., 2020). Elevations in markers of systemic inflammation such as ferritin, C-reactive protein (CRP), D-dimer, and interleukin-6 were seen, a constellation of findings described as ‘cytokine storm’(Huang et al., 2020). The cytokine storm observed in COVID-19 infection has similarities to the patterns of immune dysregulation observed in hemophagocytic lymphohistiocytosis (HLH); a disease characterized by excessive immune activation with laboratory and clinical evidence of inflammation(Opoka-Winiarska et al., 2020). An early autopsy series of COVID-19 patients from Boston Medical Center showed that three of 4 cases had histologic evidence of profound hemophagocytosis; several of these patients qualified for a formal diagnosis of secondary HLH based upon H-score(Prilutskiy et al., 2020).

Treatments for HLH are given with the intent of decreasing the inflammatory response and include options such as etoposide, an inhibitor of Topoisomerase II. Etoposide is used to selectively ablate activated T cells to control the immunoregulatory disorder in HLH(Johnson et al., 2014). Etoposide impacts T cells directly and has a broader spectrum of cytokine modulation than tocilizumab or anakinra, which target a single cytokine. (Johnson et al., 2014) Support for the use of etoposide in the treatment of HLH in adults is based upon the results of HLH-94, a large prospective pediatric study. In that study investigators used a regimen that included an 8 week induction with dexamethasone and etoposide(Henter et al., 2002). This regimen allowed the majority of patients to survive until bone marrow transplantation. In a retrospective multicenter study of 162 patients with HLH looking at predictors of mortality, use of etoposide was associated with improved survival(Arca et al., 2015). Etoposide has been previously used as salvage therapy after anti-cytokine therapy for patients with severe avian Influenza A and COVID-19 (Henter et al., 2006; Patel et al., 2021) (Patel et al., 2021). This clinical trial explored the utility of etoposide to alleviate cytokine storm and improve lung and multi-organ dysfunction in patients with life threatening COVID-19.

## Methods

### Trial Design and Randomization

This was designed as a randomized, open label, single center phase II clinical trial to evaluate the safety and efficacy of etoposide in patients with COVID-19 infection and evidence of cytokine storm. From May 15, 2020 until January 22, 2021, we enrolled hospitalized adults (>18 years of age) with COVID-19 infection as confirmed by PCR that required intubation due to COVID-19 related respiratory illness. Eligibility criteria required evidence of cytokine storm as defined by a peak ferritin >10,000 ng/mL or, peak ferritin >500 ng/mL with one or more of the following: LDH >500 U/L, D-dimer >1000 ng/mL, CRP > 100 mg/L or white blood cells (WBC) >15 k/uL. Patients were excluded if they were pregnant or breastfeeding, had a creatinine clearance <15 mL/min, bilirubin >3 mg/dL, AST or ALT > 5 times the upper limit of normal, platelets <50,000/mm^3^ or required ECMO. An amendment to the protocol allowed a single vasopressor. Enrollment targeted patients who had either failed or were considered ineligible for anti-cytokine therapy. Previous biologic therapy was allowed after a three half-life wash out period. The study protocol was approved by the Food and Drug Administration as an Investigational New Drug (IND) and by the Institutional Review Board at Boston Medical Center. It was supervised for safety by an independent medical monitor and registered with clinicaltrials.gov. Informed consent was obtained from the patient or the legally authorized representative.

Eligible patients were randomized in a 3:1 allocation ratio to receive Etoposide administered intravenously at a dose of 150 mg/m2 on Days 1 and 4 or standard of care treatment. Dose reductions of 50% were used for total bilirubin 1.5-3/0 mg/dL, AST > 2.5 times the upper limit of normal. The dose was reduced 30% if albumin was < 3.0 g/dL and 25% if creatinine clearance was 15-50 mL/min. Standard of care treatment evolved considerably while the study was open. In addition to the etoposide, therapies such as dexamethasone and remdesivir, and interventions such as prone ventilation were allowed at the discretion of the treating clinician.

### Evaluations

The patient’s clinical status was assessed at screening, day 1 prior to treatment, days 4 and 7, and at discharge or death. For patients randomized to standard of care, the day of randomization was considered day 0 of treatment. No placebo or blinding was used in this study.

### Outcome Measures

The primary outcome was improvement in pulmonary status by two categories on an 8 point ordinal scale of respiratory function. Secondary outcomes included overall survival at 30 days, cumulative incidence of grade 2 through 4 adverse events during hospitalization, duration of hospitalization, duration of ventilation and improvement in oxygenation or paO2/FIO2 ratio.

Improvement in inflammatory markers associated with cytokine storm (ferritin, CRP, d-dimer) was assessed at day 30 as were WBC and platelet count nadir and at day 30. In cases where patients were discharged or died before day 30, or if the day 30 value was missing the last lab value checked was used for analysis.

8-Point Ordinal Scale of Respiratory Function:

8 - Death;
7 - Ventilation in addition to extracorporeal membrane oxygen (ECMO), continuous renal replacement therapy (CRRT), or need for vasopressors (dopamine ≥5 μg/kg/min OR epinephrine ≥0.1 μg/kg/min OR norepinephrine ≥0.1 μg/kg/min)
6 - Intubation and mechanical ventilation
5 - Non-invasive mechanical ventilation (NIV) or high-flow oxygen
4 - Hospitalized, requiring oxygen by mask or nasal prongs
3 - Hospitalization without oxygen supplementation
2 - Discharged from hospital either to home with supplemental oxygen OR to inpatient rehabilitation/skilled nursing facility(+/-supplemental oxygen);
1 - Discharged to home without supplemental oxygen

### Statistical Analysis

The study originally planned to enroll 32 patients with a 3:1 allocation ratio. If fully accrued and assuming a 30% response rate in the control group for the primary outcome of a 2 point improvement in respiratory status on an 8 point scale at Day 14, we would detect a 50% increase in response in the Etoposide group (80% response rate in the Etoposide group) with a 2-sided α of 0.05 and 80% power. This study as originally designed was not powered to detect the minimal clinically meaningful difference in outcome, rather it hoped to provide initial signal of efficacy if improvement in response was observed.

The study was terminated early. Patients who did not receive etoposide (including the patient randomized to intervention who did not receive the etoposide) were included in the standard of care group in this modified intention to treat analysis. Only the patients who received etoposide were evaluable for toxicity. A Fisher’s exact test was used to determine if there was a significant association between receiving etoposide and dichotomous variables (2 point improvement in the ordinal scale and overall survival at 30 days). A Wilcoxon rank sum test was used to determine significance of the secondary outcomes. NCSS Statistical Software was used for analysis and (α=0.05) was used for the level of statistical significance.

## Results

### Patients

A total of 13 patients underwent screening. Of these patients, eight gave informed consent and underwent randomization. All patients were on mechanical ventilation and had received prior therapies as detailed in table 1. Several patients enrolled had already been on a ventilator for many days (median 6.6, range 1-17 days). Only one patient was randomized to standard of care. One patient was assigned to treatment but did not receive etoposide because they progressed to hemodialysis after randomization but prior to treatment. The six patients randomized to treatment received both doses of etoposide. Five of the six patients (83%) who received etoposide required dose reductions for abnormal liver function or decrease albumin. (Table 1 and Table 3)

**Table 1:** Baseline Characteristics

| Subject Number | Approximate Age (yrs) | Gender | Prior Treatment | Peak Ferritin | D-Dimer, add time (D1 or screen) | CRP | Days from intubation to consent or treatment | Dose Reduction based on enrollment labs |
| --- | --- | --- | --- | --- | --- | --- | --- | --- |
| 1 | 51-55 | Male | Anakinra | 14038 | 852 | 443.4 | 11 | None |
| 2 | 61-65 | Female | Sarilumab, Dexamethasone | 4553 | 1201 | 309.1 | 17 | 50%: LFT |
| 3 | 61-65 | Male | Remdesivir, Dexamethasone | 5086 | 989 | 242.7 | 4 | 50%: LFT & albumin |
| 4 | 71-75 | Male | Dexamethasone | 5441 | 1162 | 223.7 | NA- get the number |  |
| 5 | 51-55 | Male | Dexamethasone | 3486 | 2445 | 138.1 | 4 | 30%: albumin |
| 6 | 75-80 | Female | Dexamethasone | 513 | 279 | 68.6 | NA- get the number |  |
| 7 | 85-90 | Male | Dexamethasone, Ivermectin, Remdesivir | 1053 | 281 | 56.9 | 3 | 30%: albumin |
| 8 | 75-70 | Male | Dexamethasone, Remdesivir | 1753 | 671 | 74.3 | 6 | 30%: albumin |

### Primary Outcome

Two of 6 (33%) patients randomized to receive etoposide met the primary endpoint of improvement of two points on the respiratory ordinal scale. One patient was discharged to rehab with a tracheostomy requiring ongoing ventilator support so did not meet the primary outcome measure but ultimately recovered. The patient randomized to treatment who did not receive etoposide due to rapid decline in renal function died 20 days after enrollment. The one patient randomized to standard of care was discharged on room air at day 28, thereby meeting the primary endpoint (50%) Three of 6 patients (50%) treated with etoposide died on the ventilator. The cause of death was hypoxemic respiratory failure from COVID lung injury in all cases. There was no significant association between receiving etoposide and a 2 point improvement on the ordinal respiratory scale (2 of 6 etoposide patients (33%) vs 1 of 2 (50%) SOC patients, p>0.999, Fisher’s exact test) (Table 2)

**Table 2.** Results:

| Subject Number | Assigned Treatment | Score at enrollment | Score at Discharge | Outcome at Discharge | Days from treatment/ enrollment to extubation | Days from treatment/ enrollment to discharge or death |
| --- | --- | --- | --- | --- | --- | --- |
| 1 | Etoposide | 6 | 1 | Home on Room Air | 12 | 22 |
| 2 | Etoposide | 6 | 6 | Rehabilitation with a tracheostomy & weaning ventilator support | More than 30 | 38 |
| 3 | Etoposide | 6 | 8 | Deceased |  | 8 |
| 4 | Etoposide, never received (analyzed as SOC) | 6 | 8 | Deceased |  | 20 |
| 5 | Etoposide | 6 | 1 | Home on Room Air | 5 | 11 |
| 6 | Standard of Care | 6 | 1 | Home on Room Air | 15 | 28 |
| 7 | Etoposide | 6 | 8 | Deceased |  | 11 |
| 8 | Etoposide | 6 | 8 | Deceased |  | 12 |

| Subject number | Dose Reduction | Adverse Events |
| --- | --- | --- |
| 1 | None | Grade 1: Anemia, Leukopenia |
| 2 | 50%: baseline LFT<br>Day 4 30% for Albumin <3 | Grade 2 Anemia<br>Worsening renal function |
| 3 | 50%: baseline LFT & albumin | Grade 1 Transaminitis,<br>Grade 3 Thrombocytopenia<br>Grade 2 Anemia<br>Grade 4 Neutropenia<br>Neutropenic Fever |
| 4 | NA | NA |
| 5 | 30%: baseline albumin | None |
| 6 | NA | NA |
| 7 | 30%: baseline albumin | Ventilator associated pneumonia (not neutropenic) |
| 8 | 30%: baseline albumin | Grade 4 Anemia<br>Grade 4 Neutropenia<br>Grade 4 Thrombocytopenia |

### Secondary Outcomes

There was no significant association between receiving etoposide and survival at day 30 in a modified intention to treat analysis, (3 of 6 (50%) for etoposide, 1 of 2 (50%) for SOC p>0.999; Fisher’s exact test). Likewise, there were no significant associations between change in ferritin (1235 ng/L etoposide vs -1771 ng/L p = 0.14), CRP (−49mg/L etoposide vs -74 mg/L SOC, p = 0.52) or d-dimer (480 ng/mL etoposide vs -2243 ng/mL SOC, p = 0.64) from pretreatment levels until day 30 (or discharge or death) levels.

### Safety and Adverse events

Despite dose reductions, four patients of 6 (66%) experienced bone marrow suppression during the study period related to etoposide, with two patients (33%) experiencing grade 4 neutropenia and grade 3 or 4 thrombocytopenia. One patient experienced febrile neutropenia requiring broad spectrum antibiotics. Three patients (50%) died or had care withdrawn while still in their hematologic nadir although none of these deaths were from a complication of myelosuppression (new infection or bleeding). One patient developed grade 1 AST/ALT elevation and one patient had grade 4 acute kidney injury requiring dialysis.

## Discussion

Etoposide is useful to control the dire immune dysregulation seen in hemophagocytic lymphohistiocytosis. The similiarities between HLH and the pathologic inflammation seen in severe COVID suggested that it may also be helpful to blunt the impact of cytokine storm from COVID. This trial attempted to evaluate the utility of etoposide in critically-ill patients with severe COVID requiring mechanical ventilation.

Accrual to this study was halted early due to the rapidly changing demographics of the infection and the emergence of other available treatment options. The limited sample size hampers interpretation. From the available data, inconsistent myelosuppression is the major adverse effect of etoposide for COVID, despite the use of dose reductions. This toxicity will limit etoposide’s applicability in critically ill patients with COVID. Limitations to this study include the small sample size and the rapid changes in standard of care for COVID that occurred during the time of this study. It is also acknowledged that the intervention for many of these patients occurred after the initial period of cytokine storm had resolved. Within these significant limitations, there was no clear trend in improvement in respiratory function or secondary outcome measure that supports further exploration in this population.

## Data Availability

All data produced in the present study are available upon reasonable request to the authors

## Notes

### Competing Interest Statement

The authors have declared no competing interest.

### Clinical Trial

Results Record H-40102 (NCT04356690)

### Funding Statement

Boston University Clinical & Translational Science Institute NIH identifier 1UL1TR001430.

### Author Declarations

IRB of Boston Medical Center gave ethical approval for this work

